# Effect of physical activity calorie equivalent (PACE) labels on energy purchased in cafeterias: a stepped-wedge randomised controlled trial

**DOI:** 10.1101/2022.02.26.22271547

**Authors:** James P. Reynolds, Minna Ventsel, Alice Hobson, Mark A. Pilling, Rachel Pechey, Susan A. Jebb, Gareth J. Hollands, Theresa M. Marteau

## Abstract

**Background:** A recent meta-analysis suggested that using physical activity calorie equivalent (PACE) labels results in people selecting and consuming less energy. Only one included study was conducted in a naturalistic setting, in four convenience stores. The current study aimed to estimate the effect of PACE labels on energy purchased in worksite cafeterias.

**Methods and findings:** A stepped-wedge randomised controlled trial to test the effect of PACE labels (which include kcal content and minutes of walking required to expend the energy content of the labelled food) on energy purchased. The setting was ten worksite cafeterias in England, which were randomised to the order in which they introduced PACE labels on selected food and drinks following a baseline period. The study ran for 12 weeks with over 250,000 transactions recorded on electronic tills. The primary outcome was total energy (kcal) purchased from intervention items per day. The secondary outcomes were: energy purchased from non-intervention items per day, total energy purchased per day, and revenue. Regression models showed no evidence of an overall effect on energy purchased from intervention items, -1.3% (95% CI -3.5% to 0.9%) during the intervention. Of the 10 cafeterias, there were null results in five, significant reductions in four, and a significant increase in one. There was also no evidence for an effect on energy purchased from non-intervention items, -0.0% (95% CI -1.8% to 1.8%), and no clear evidence for total items -1.6% (95% CI -3.3% to 0.0%). Revenue increased during the intervention, 1.1% (95% CI 0.4% to 1.9%). Study limitations include using energy purchased and not energy consumed, and access only to transaction-level sales, rather than individual-level data.

**Conclusion:** Overall, the evidence was consistent with PACE labels not changing energy purchased in worksite cafeterias. There was considerable variation in effects between cafeterias, suggesting potentially important unmeasured moderators.

**Trial registration:** The study was prospectively registered on ISRCTN (date: 30.03.21; ISRCTN31315776).

## Introduction

Excess energy intake contributes to over 60% of the UK adult population being overweight or obese, contributing to high and rising levels of type 2 diabetes and 13 different types of cancer (GBD Obesity Collaborators, 2017; Steel et al., 2018; Swinburn et al., 2009). Interventions to reduce overconsumption of energy must form a central part of wider strategies to tackle overweight and obesity.

One approach to reducing excess energy intake has been to add labels on food and drinks to inform people about the energy content of the product. A Cochrane systematic review and meta-analysis of three nutritional labelling studies in restaurants suggested a reduction in energy purchased by 47kcals per meal (Crockett et al., 2018) whereas a separate meta-analysis on six labelling studies in restaurants concluded that there was no effect on energy ordered (Long et al., 2015). However, the quantity and quality of the available evidence is limited. Two randomised trials in worksite cafeterias that were published after these reviews provided no evidence for an effect of simple energy labelling (kcal) on energy purchased (Vasiljevic et al., 2018, 2019).

An alternative to labelling the energy content is to convert this information into the physical activity needed to expend the energy in that product. PACE (Physical Activity Calorie Equivalent) labels typically include the energy content, the equivalent energy in terms of physical activity and an image representing the type of physical activity – usually walking or running. A recent systematic review concluded that PACE labels may reduce energy selected from menus and decrease the energy consumed when compared to no labelling or other types of labelling such as kcal labelling (Daley et al., 2019). However, of the 15 included studies, most were of an unclear risk of bias and only one was conducted in a naturalistic setting (Bleich et al., 2011). The remaining 14 studies were conducted online (n = 8) or in non-naturalistic settings (n = 6), and recent reviews of labelling studies suggest that effects are typically largest in online studies and smallest in naturalistic settings (Clarke et al., 2021; Long et al., 2015). The one naturalistic study (Bleich et al., 2011) investigated the effect of PACE labels on sugar-sweetened beverages in four convenience shops in the USA. The results suggested that participants were less likely to purchase a sugar sweetened beverage when PACE labelling was added (OR = 0.51). A further naturalistic study in three worksite cafeterias, published after the review, found that PACE labels resulted in a significant decrease in energy purchased of approximately 40 kcals per meal (Viera et al., 2019). This study recruited participants who regularly used the cafeterias and asked them to photograph their food at multiple points over time. This study therefore only includes a subset of the total customers and a subset of the total purchases that these participants made over the study period. Based on the quantity and quality of evidence, considerable uncertainty remains about the effect of PACE labels to reduce energy purchased and consumed, warranting further studies at low risk of bias, and conducted in real world settings.

The aim of the current study is to estimate the effect of PACE labels on energy purchased in worksite cafeterias. The limitations of existing naturalistic studies are addressed in three ways: by implementing the intervention in cafeteria settings across a wide range of food and drinks; collecting sales data from electronic tills to ensure the availability of data on every purchase made by every customer of the cafeterias throughout the study period; and, by conducting the study in a larger number of sites to increase the study power and test the generalisability of the main effects to multiple cafeterias. The study is designed to test the hypothesis that customers purchase less energy when food and drinks feature PACE labels.

## Methods

The study was prospectively registered on ISRCTN (ISRCTN31315776) and a detailed analysis plan was uploaded to the Open Science Framework (https://osf.io/2a5cg/?view_only=95d3d6a38cf047588f4e8365207ef1f4) during data collection, but before data cleaning or analysis had commenced. The CONSORT extension checklist for stepped-wedge trials is attached as a supplement (see “S1_CONSORT_Checklist”). The Cambridge Psychology Research Ethics Committee based at the University of Cambridge approved the trial on 08.12.20 (No. PRE.2020.105). The research team obtained informed and written consent from a representative of the catering company on behalf of the participating cafeterias.

### Cafeterias

Ten worksite cafeterias were recruited through a major UK catering company (see Fig 1) and were based within worksites belonging to different companies. There were three eligibility criteria for participation: *i*. located in Great Britain, *ii*. at least 500 employees based at the cafeteria, and *iii*. sales data are recorded using electronic point-of-sale tills. Twenty cafeterias were screened for eligibility and 10 participated in the study (Fig 1). The remaining 10 cafeterias were not eligible due to violating the second eligibility criterion. At recruitment, participating sites employed between 500 and 7200 staff. The smallest cafeteria, Cafeteria 8 (230 employees), had fewer employees by the end of data collection than was reported during recruitment due to covid-related staffing changes (see Table 1).

**Table 1.** Demographic characteristics of employees in participating sites

| Cafeterias | Number of employees | Male (%) | Age (mean) | Typical job roles | Full time (%) |
| --- | --- | --- | --- | --- | --- |
| Total | 19361 | 72 | 40 | - | 77 |
| 1 | 1800 | 70 | 37 | Pilots/ Office/ Admin/ Facilities/ Management | 80 |
| 2 | 697 | 75 | 47 | Processing/ Drivers Office Staff | 62 |
| 3 | 1500 | 80 | 44 | Engineers/ Designers/ Accountants/ Facilities/ Management | 100 |
| 4 | 2361 | 80 | 26 | - | - |
| 5 | 850 | 70 | 45 | Processing/ Drivers Office Staff | 60 |
| 6 | 3473 | 95 | 39 | Manufacturing associates/Admin / Management | 97 |
| 7 | 500 | 60 | 47 | Processing/ Drivers Office Staff | - |
| 8 | 230 | 50 | 30 | Manufacturing/ Management & Office | 100 |
| 9 | 750 | 66 | 42 | Drivers/ Office/ Facilities/ Sales/ Admin/ Delivery/ Managers | 40 |
| 10 | 7200 | - | - | - | - |

*Note.* Some data are missing as the cafeterias managers did not provide it.

**Fig 1.**
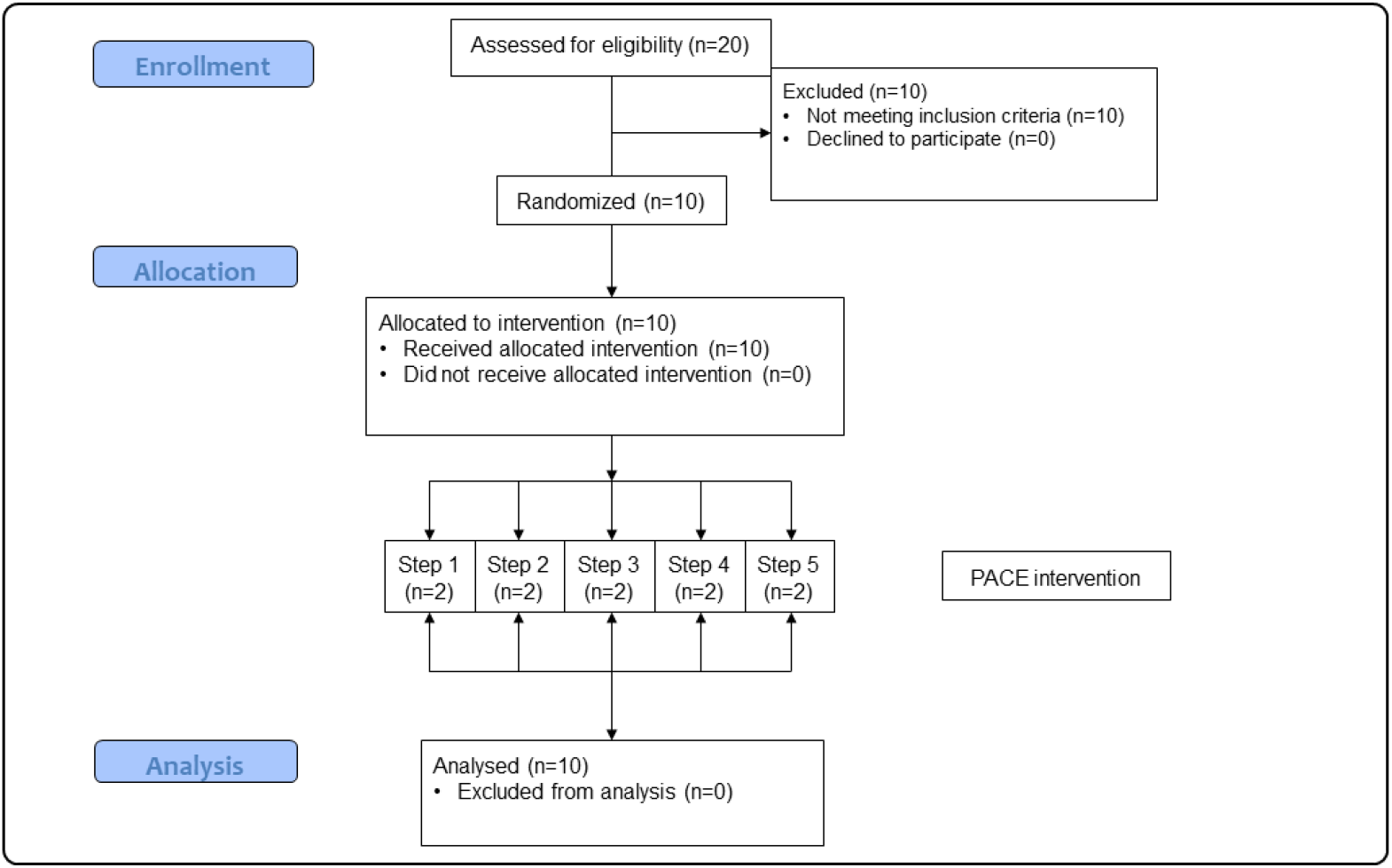
CONSORT flow diagram *Note. n* refers to cafeterias. Each step refers to the different weekly periods in which cafeterias start the interventions. See Fig 3 for the timings of these steps.

The recruitment strategy was based on practical limitations, specifically, the maximum number of eligible cafeterias that we could recruit from our collaborating company. The stepped wedge design was used for pragmatic reasons, as they are typically preferred to a parallel groups randomised controlled trial (RCT) when study resources only allow a staggered implementation of the intervention(s) (Reynolds et al., 2021).

### Study periods

#### Baseline

Baseline was a period of business-as-usual for the cafeterias when sales data were collected before the intervention period. During baseline, most pre-existing labels and menus only featured the product name and price. There were some standardised front of pack nutrition labels on branded products (e.g., Coca-Cola) and in-house products (e.g., muffins) on which the energy content was provided in small print.

There were largely no energy labels on shelf edge labels or menus.

### Intervention

The PACE label intervention comprised adding two new pieces of information to the product: the energy content of the product and the physical activity calorie equivalent of this value, expressed in the minutes of walking that would be needed to expend the energy in the product (see Fig 2). These labels were co-produced by the research team, the catering staff, and the catering managers. In the typology of interventions in proximal physical micro-environments (TIPPME) (Hollands et al., 2017), this is classified as an *Information x Product* intervention. These labels were added in up to four locations at each cafeteria: *i*. shelf-edge labels, *ii*. menus next to food and drink displays, *iii*. individual tent cards next to food and drink displays, and *iv:* on stickers that were attached to the product packaging. Posters that explained the meaning of the labels were also put up at participating cafeterias and the service staff were briefed in case customers asked them any questions.

**Fig 2.**
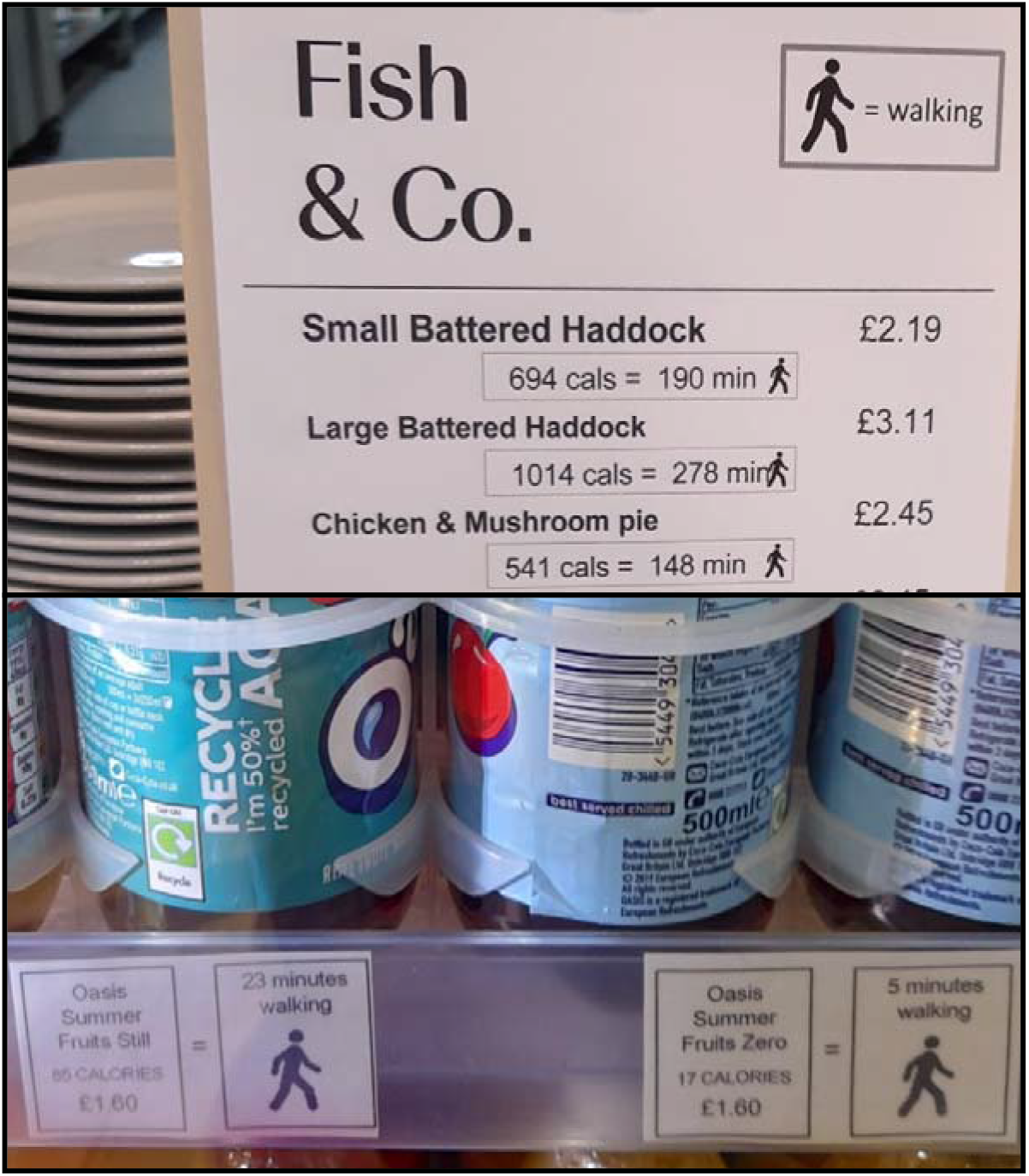
Example menu (top) and shelf-edge labels (below) used during the PACE intervention

### Study design

A stepped-wedge design was used across a period of 12 weeks (06 April 21 to 28 June 21). The 10 cafeterias were randomly allocated to the week in which the intervention was implemented (see Fig 3). The baseline period lasted between four and eight weeks. Weeks 1 to 4 comprised the minimum baseline period. From week 5 until week 9, two cafeterias a week implemented the PACE label intervention, which lasted until the end of week 12, when the study ended.

**Fig 3.**
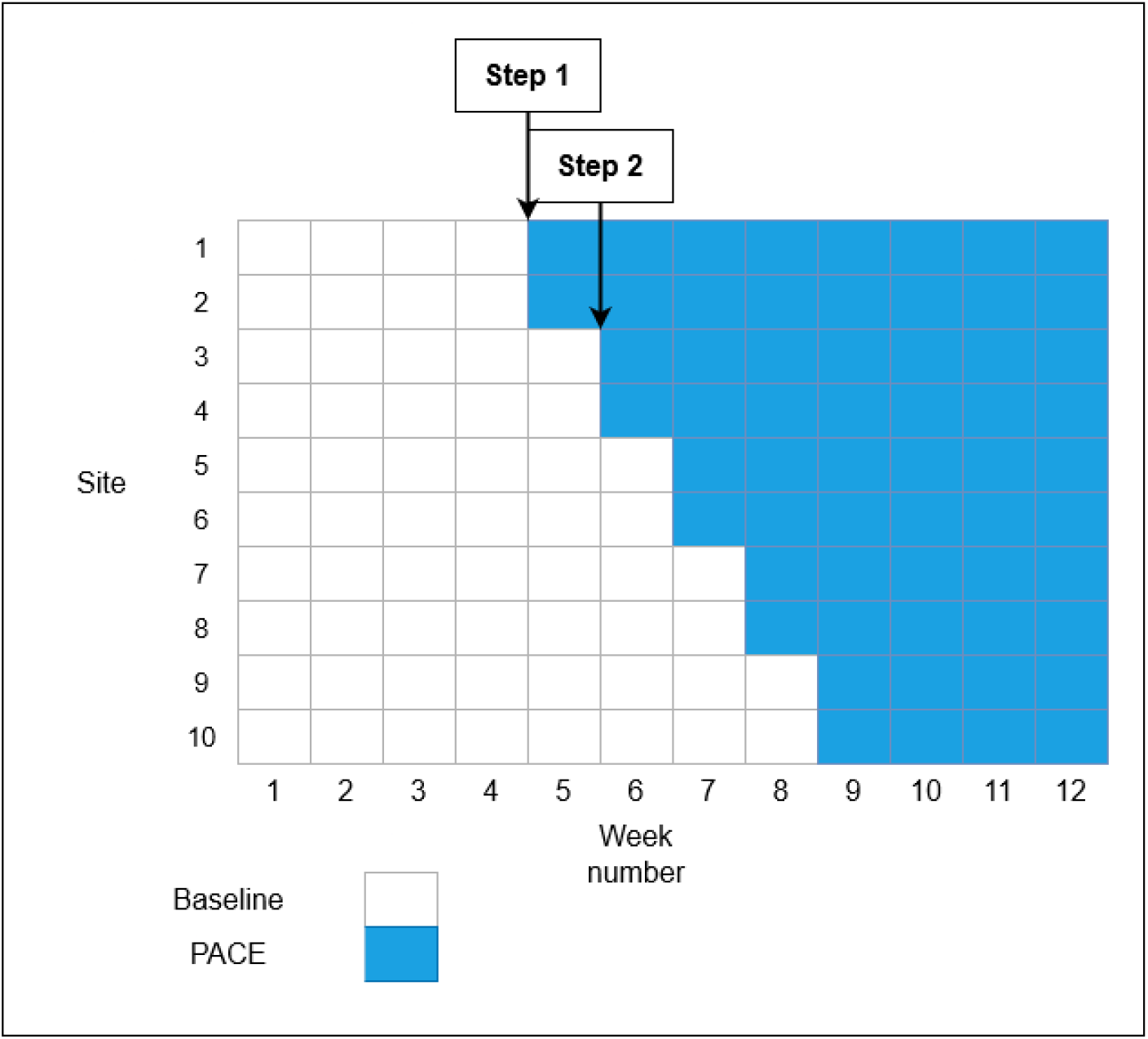
Stepped-wedge study design

### Randomisation and blinding

Participating cafeterias were randomly allocated to the time at which the interventions were implemented. The randomisation was performed by a Statistician who was blinded to the identity of the cafeterias. The Statistician allocated a list of anonymised cafeteria names using the rank of random numbers from Excel. Staff at each cafeteria were told not to inform customers that the labels were part of research. In response to questions from customers about the labels staff were instructed to say they were being trialled as part of a health initiative. Staff and customers could not be blinded to the interventions as the labels are designed to be seen and read by customers. Cafeterias were informed about their allocation (the week in which they were to implement the interventions) after recruitment and before data collection, which allowed time for them to prepare for the interventions.

### Procedure

#### Covid-19

The current study took place during the Covid-19 pandemic which affected the fidelity checks. The original plan was to visit the cafeterias in person, but the initial visits were replaced with video call interviews with cafeteria managers and the regular site visits were replaced with photos of the food and drink taken by members of staff every week.

#### Implementation

The PACE labelling intervention was implemented by catering staff and managers at each cafeteria following training and assistance from senior management within the catering companies and the research team. To maximise coverage, the catering staff were instructed to add labels to all products on sale for which the energy content was known.

#### Fidelity

Detailed photos of the products on sale and their labels were requested to be sent once a week to the research team for checks. During the baseline period these checks were to ensure that *i*. no PACE labels were present, and *ii*. no energy (kcal) labels were present. During the PACE labelling period, these checks were to ensure that *i*. the PACE labels were present, and *ii*. the energy (kcal) and PACE values on these new labels were accurate.

Any violations to these criteria were reported to a manager responsible for the cafeteria with a request to rectify the violation and provide photographic evidence of the rectification within 24 hours. Adherence to or violation of the planned implementation was recorded for use in secondary analyses.

### Measures

All outcomes were calculated using data from the electronic point of sale tills that were used at each cafeteria. Data were collected during every day that the cafeteria was open during the length of the study.

#### Primary outcome

This comprised energy (kcal) purchased from intervention items per day. This was calculated using the total number of sales for all items that featured a PACE label and the energy content for each of these items.

To determine which products featured a PACE label we used the digital copies of the PACE labels and menus. These digital copies were Word and PowerPoint documents that contained every PACE label and menu. These were printed off at each cafeteria and then added to the products on sale.

Energy content (kcals) was available for most products (97%) on sale at the cafeterias. This information was obtained from the catering provider, the cafeterias, and by searching online. For a further 16 products (1%) energy content was estimated by taking the average from three similar products, resulting in energy content for 98% of all products. For the remaining 2% it was not possible to reliably estimate energy content using any approach and therefore these data were not included in the analysis.

#### Secondary outcomes

1. Total energy (kcal) purchased per day from non-intervention items (products that did not include PACE labels).
2. Total energy (kcal) purchased from all food and drink products. This included all products, including intervention and non-intervention items.
3. Total revenue from each cafeteria. This was calculated from the total number of items sold in all categories and the price of each item.

#### Covariates

1. Total number of transactions: the number of distinct payments to purchase products in the cafeteria, as a proxy measure for the number of customers per day
2. Time: day number of the trial at each cafeteria, starting from 1 within Baseline or Intervention periods, common to all cafeterias
3. Time (week of trial): common to all cafeterias, fitted as a random effect
4. Time (day of the week)

### Analyses

Generalised additive linear mixed models (Stasinopoulos et al., 2017) were used to estimate the overall potential impact of the PACE intervention compared to baseline due to markedly different variability (heteroscedasticity) at cafeterias. Cafeterias were fitted as random effects, with the effect of the day of the week allowed to vary by cafeteria as a random nested term due to regular weekly patterns at each cafeteria. The effect of week of the trial was fitted as a random factor common to all cafeterias, due to weekly changes observed in the data. To allow for potential linear time trends, the day number of the trial was fitted as a continuous fixed effect. Due to the irregular and rare instances of Cafeteria 3 opening at weekends, these three data points were removed as they were insufficient for parameters to be estimated.

When estimating the intervention effect at each of the ten cafeterias, a Bonferroni adjustment for multiple testing was applied where the threshold for significance was 5%/10 and 99.5% confidence intervals were presented. Model diagnostics were assessed using variance inflation factors, residual plots, quantile– quantile plots, worm plots and correlation function plots for the additive models; these diagnostics were acceptable. Exploratory plots of weekly-aggregate data were also examined, and heterogeneity was still present.

There were two prespecified sensitivity analyses. First, during data collection researchers asked till staff at the cafeterias which if any buttons or barcodes were not working, meaning that certain products were mis-sold under another product’s button or barcode. The primary analysis adjusts for this error by taking the average energy content for the multiple products that were sold under a specific till button whereas the first sensitivity analysis reports the results without this adjustment. Second, Cafeteria 10 provided photographic evidence of label implementation on week 12 instead of the planned week 9. This second sensitivity analysis removes this cafeteria’s data from weeks 9, 10, and 11, to account for the possibility that the labels were not implemented. The primary analysis assumes that labels were implemented as intended and does not remove these data.

## Results

### Implementation

All cafeterias reported implementing the labels on the planned week, with 9/10 cafeterias providing immediate photographic evidence of this implementation.

The fidelity checks suggested that the PACE labelling intervention was applied to 93% of products on sale (Table 2). The most common locations for the PACE labels to be added was on shelf-edges (10/10 cafeterias) and menus (9/10 cafeterias).

**Table 2.** Implementation of PACE labelling overall and by cafeteria

| Cafeteria | Proportion (%) of total products that received PACE labels | Location: Shelf edge labels | Location: Menus | Location: Tent cards | Location: Stickers on products |
| --- | --- | --- | --- | --- | --- |
| **Overall** | 93 | 10/10 | 9/10 | 5/10 | 1/10 |
| **1** | 97 | Yes | Yes | No | No |
| **2** | 100 | Yes | Yes | No | No |
| **3** | 92 | Yes | Yes | No | No |
| **4** | 93 | Yes | Yes | No | No |
| **5** | 95 | Yes | Yes | Yes | No |
| **6** | 96 | Yes | Yes | Yes | No |
| **7** | 90 | Yes | Yes | Yes | No |
| **8** | 92 | Yes | Yes | Yes | Yes |
| **9** | 84 | Yes | Yes | Yes | No |
| **10** | 90 | Yes | No | Yes | No |

### Primary outcome

There was no evidence that the PACE intervention resulted in an overall change in energy purchased from intervention items, -1.3% (95% CI -3.5% to 0.9%), *p* = .236. This effect size is equivalent to -5kcals per transaction (95% CI -12kcals to 3kcals). These results are shown in Fig 4 and the unadjusted data are shown in Table 3.

**Table 3.** Unadjusted data (mean [sd]) for daily purchases, revenue, and prices in cafeterias during intervention periods

|  | Baseline | PACE |
| --- | --- | --- |
| Energy (kcal) purchased, per cafeteria from intervention categories | 148350 [87637] | 163949 [97267] |
| Number of transactions, per cafeteria | 315 [187] | 365 [204] |
| Revenue (£), per cafeteria | 979 [631] | 1166 [716] |

**Fig 4.**
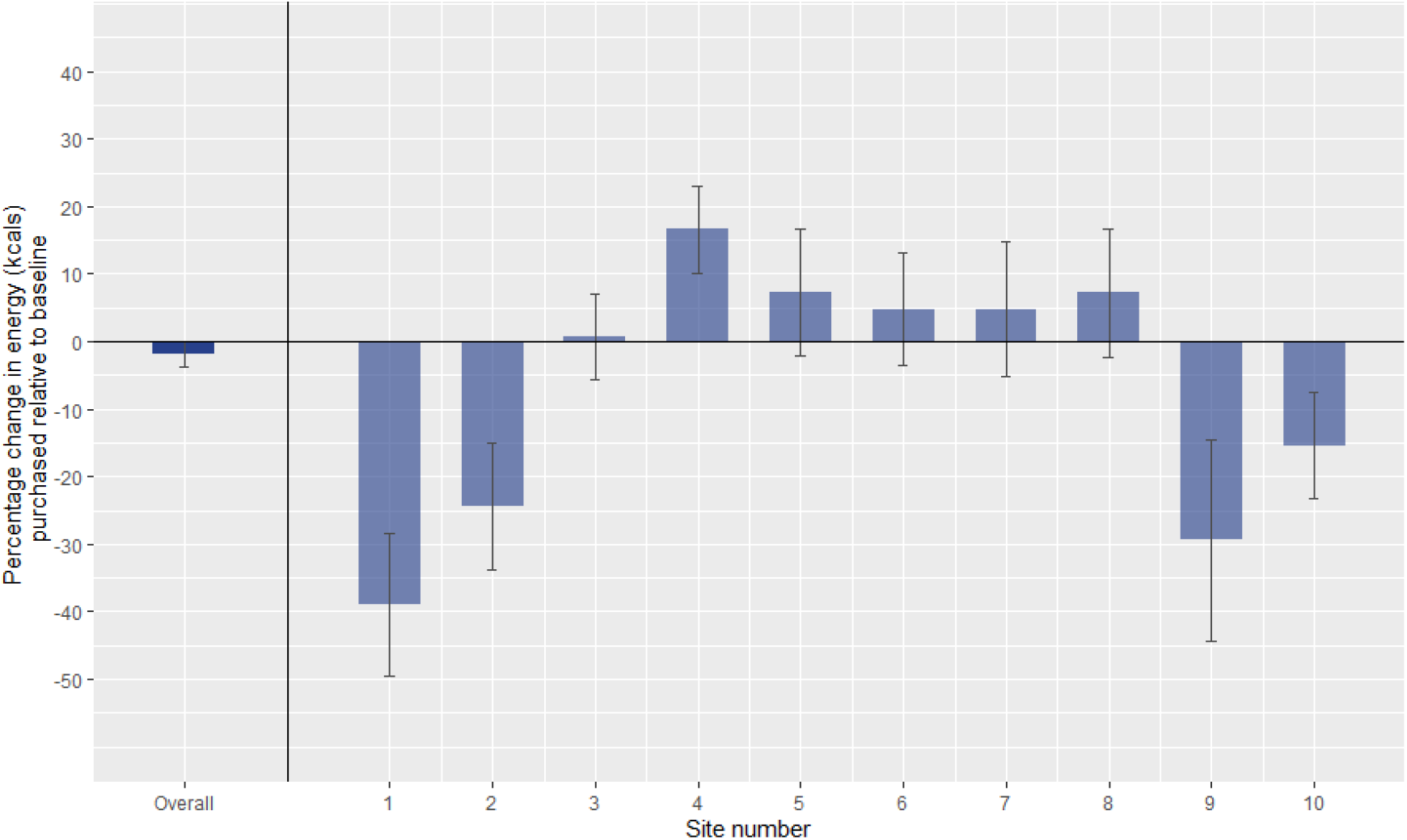
The effects PACE on energy (kcals) purchased per day relative to baseline. *Error bars represent 95% confidence intervals for the overall effects and 99*.*5% confidence intervals (Bonferroni adjustment) for the by-cafeteria effects*.

#### Individual cafeteria effects

There was no evidence that the PACE intervention resulted in a change in five cafeterias: cafeteria 3: - 0.8% [99.5% CI -5.6% to 7.1%]; cafeteria 5: 7.2% [99.5% CI -2.2% to 16.7%]; cafeteria 6: 4.7% [99.5% CI -3.6% to 13.1%]; cafeteria 7: 4.8% [99.5% CI -5.1% to 14.7%]; and cafeteria 8: 7.3% [99.5% CI - 2.3% to 16.8%].

The PACE intervention significantly reduced energy purchased in four cafeterias: cafeteria 1: -38.9% [99.5% CI -49.6% to -28.3%]; cafeteria 2: -24.5% [99.5% CI -33.8% to -15.1%]; cafeteria 9: -29.4% [99.5% CI -44.3% to -14.6%], and cafeteria 10: -15.4% [99.5% CI -23.3% to -7.6%].

In one cafeteria the PACE intervention significantly increased energy purchased: cafeteria 4: 16.6% [99.5% CI 10.2% to 23.0%]).

### Secondary outcomes

#### Energy purchased from non-intervention items

There was no evidence that the PACE intervention resulted in a change in energy purchased from non-intervention items, -0.0% (95% CI -1.8% to 1.8%), *p* = .986.

#### Total energy purchased (intervention and non-intervention items)

There was no clear evidence that PACE intervention resulted in a change in total energy purchased from all products, -1.6% (95% CI -3.3% to 0.0%), *p* = .051, albeit with the upper CI estimate touching zero indicating that it is more likely that this intervention decreased total energy purchased from all products.

#### Revenue

During the PACE intervention there was a significant increase in revenue for the cafeterias, 1.1% (95% CI 0.4% to 1.9%), *p* = .002. This effect size is equivalent to an additional £11.06 per day per cafeteria, or an additional £0.03 per transaction.

#### Sensitivity analyses

The two pre-specified sensitivity analyses were consistent with the primary results. First – for the analysis in which the energy estimates were not adjusted for incorrect button presses – there was no evidence that the PACE intervention resulted in an overall change in energy purchased from intervention items, -1.3% (95% CI -3.0% to 0.4%), *p* = .132. Second – for the analysis in which we remove some data from the cafeteria that did not provide timely evidence of intervention implementation – there was no evidence that the PACE intervention resulted in an overall change in energy purchased from intervention items, -0.8% (95% CI -3.0% to 1.5%), *p* = .503.

#### Data checks

Results from the main analysis for some of the cafeterias produced larger effect sizes than were predicted. This includes the PACE intervention resulting in a 39% drop (−50% to -28%) in energy purchased at Cafeteria 1 and a 29% drop (−44% to -15%) at Cafeteria 9. To provide reassurance about the accuracy of the data collection, we conducted a series of checks to validate the accuracy of these findings, all of which substantiated these findings. First, we checked recording validity at the till sales level to exclude the possibility that a single transaction had erroneously been recorded multiple times (e.g., 50 fish and chips in a single transaction, say). Second, we checked for outliers (defined in the preregistration protocol as three units using median absolute deviation) at the aggregate level (daily & weekly) in which energy purchased per cafeteria was examined. Third, we conducted four sensitivity analyses on data at the level of the cafeteria: i. removing outliers, ii. not adjusting for till button errors (see ‘Sensitivity analysis’ section above), iii. removing data from the cafeteria that failed to provide evidence of implementation, iv. using a model that more simplistically assumes equal variance across cafeterias.

## Discussion

There was no overall evidence that PACE labels changed energy purchased when compared to baseline in ten worksite cafeterias across England. This conclusion was supported from all three indicators of energy purchased: energy purchased from products featuring a PACE label, energy purchased from products not featuring a PACE label, and total energy purchased from all food and drinks. The cafeteria-level analysis showed considerable variation in effects for the primary outcome: of the 10 cafeterias, there were null results in five, significant reductions in four, and a significant increase in one.

This is the largest study to date that evaluates the impact of PACE labels on food and drink purchases within a naturalistic setting and therefore provides the best estimate for the effectiveness of PACE labels at changing the purchasing of food and drinks. The pooled results of the current study estimate the effect of PACE labels at -1.3% (95% CI -3.5% to 0.9%) which is equivalent to an effect of -5 kcals (95% CI -12 kcals to 3 kcals) per transaction. These results do not overlap with the confidence intervals reported by Daley et al (2019): -80 kcals (95% CI -137 kcals to -24 kcals) nor is the main effect replicated. The current results are therefore not consistent with this review. The current results also appear to be inconsistent with the results of Bleich et al (2011) and Viera et al (2019). There are several possible explanations for these apparent differences in outcomes. The Daley review largely included hypothetical, online selection studies and non-naturalistic studies in which participants were recruited in university settings, and given menus with PACE labels by researchers, which may not generalise to typical behaviour in restaurants, supermarkets, or cafeterias. Previous labelling studies have suggested that effects in online studies tend to produce larger effect sizes than in lab settings, and lab settings tend to produce larger effect sizes than those observed in naturalistic settings (Clarke et al., 2021; Long et al., 2015). A study in four convenience stores (Bleich et al., 2011) found significant effects, but the target of intervention was sugar-sweetened beverages, which only made up a small proportion of the sales in the current study. It remains possible, yet untested, that PACE labels have different effects when applied to different products. A study in three cafeterias (Viera et al., 2019) relied on a subset of the cafeteria’s customers sending photos of their meals for two week periods every three months and therefore the results do not provide a comprehensive account of how the labels influenced overall customer purchases within these cafeterias. In contrast to these studies, the current study applied PACE labels to many categories of food and drink (hot meals, sandwiches, cold drinks, desserts, etc) and collected data from every sale for which energy content was available (98%), totalling over 250,000 transactions across 10 cafeterias.

In the current study we observed a marked difference in the direction and magnitude of the intervention effect in different cafeterias. This variability should sound a note of caution regarding results of trials conducted in small numbers of cafeterias. It is possible that there are contextual factors, such as the type of food and drink sold at the cafeterias, the degree of label implementation, or whether the cafeteria is used for eat-in vs “grab-and-go”, which impact on effectiveness. There may also be demographic differences in who used the different cafeterias and their responses to PACE labels which could account for some of the variation between cafeterias. Table S1 in the supplement shows that the cafeterias varied largely in terms of the food and drink that were regularly sold. For example, the proportion of energy purchased from hot meals ranged from 2% to 27%, breakfasts from 5% to 58%, sandwiches from 4% to 19%, and hot drinks from 0% to 13%. Visual inspection of Table S1 suggests that PACE labels show larger effects in cafeterias that sell more discretionary items. It is possible that PACE labels are more effective at reducing purchasing of discretionary items compared to main meals, which may explain why the effect sizes are larger at certain sites. Further research is needed to confirm this. Regardless of the underlying explanation for the variation across sites, these possible sources of variation could explain why the results here do not support the conclusions of earlier research. Namely, that the effectiveness of the PACE labels is contingent on contextual factors that differed between the average cafeteria in the current study and the settings used in previous research.

The results of the secondary analyses were consistent with the primary analysis. Namely, there was no clear evidence of an overall effect of the intervention on energy purchased from non-intervention items (i.e., products without PACE labels) or all items (i.e., non-intervention items and intervention items combined). There was evidence that revenue increased during the PACE period relative to the baseline period. As there was no detected effect of the PACE labels on energy purchased, it seems unlikely that this 1.1% increase in revenue was due to addition of the PACE labels, and could be explained by inflation, which increased throughout the Study period by 1.7% as measured using the Consumer Price Index (Rate Inflation, n.d.) and cannot be easily controlled in a stepped wedge design. Some studies that have tested different interventions in cafeterias have detected reductions in revenue (Reynolds et al., 2021), but there is no evidence that this should be a barrier to implementation of PACE labels.

### Strengths and limitations

The current study was the largest to date (to our knowledge) to implement and test the effectiveness of PACE labels. PACE labels were implemented on the majority of products in cafeterias and outcome variables were measured objectively using data derived from electronic point-of-sale tills. The main limitations of this study were that we were not able to assess consumption of the food and drink purchased, although sales data in cafeterias are normally a good proxy for consumption in these settings as only a small percentage of food is wasted by individuals (Hinton et al., 2013; Vermote et al., 2018).

These sales data were also only described at the transaction level and being able to link transactions to individuals would provide more useful information for inferring causation. Individual identifiers were not possible to acquire due to the companies’ desire not to share this data without an external organisation.

Data collection occurred between April and June 2021 in the UK, during the COVID-19 pandemic, and at a time when various government guidelines were in place. This includes stay at home restrictions during April and a phased re-opening during May and June. The 2m social distancing rule was also in place during this time. The cafeterias were still in operation during this time as they were workplace cafeterias within businesses that were exempt from closure, however it is possible that eating behaviours may have been influenced by the guidelines or wider effects of the pandemic. Although this was not tested, this could include an increased likelihood for takeaway foods from cafeterias as customers may have been less inclined to sit and eat in the cafeterias. While it is not possible to evaluate the consequences of COVID-19 virus, restrictions, and guidelines on eating behaviours in the current study, the results should be interpreted in light of these contextual factors.

### Implications for policy and practice

There is no overall evidence from this study that PACE labels would either reduce or increase the amount of energy purchased in worksite cafeterias. However, studies of other types of labelling have suggested that companies may provide a healthier food offer when labelling is present (Theis & Adams, 2019). This is difficult to test until labelling is fully implemented in a cafeteria. It is also possible that providing information on physical activity equivalence may increase activity, although the one study that has tested this prediction reported inconclusive results (Deery et al., 2019). These possible benefits could support the introduction of PACE or other labelling schemes if accompanied by a formal evaluation to test the full range of impacts. Large scale rollout would also allow for a formal evaluation of the contextual factors that may influence intervention effectiveness.

### Conclusion

The current study provides evidence consistent with PACE labels not influencing food or drink purchasing in ten cafeterias. Despite an overall null effect, there were significant effects in both directions at individual cafeterias though mostly towards a reduction due to PACE. Based on these effects, and the uncertainties around which contexts could lead to increases or decreases in energy purchased at individual cafeterias, there is insufficient evidence to justify implementing the labels in cafeteria settings. Identifying the characteristics of settings in which PACE labels could be beneficial is important if these are to be implemented more widely.

## Data Availability

This study includes sales data from the collaborating companies. Due to contractual restrictions to the use of the sales data, these data cannot be made openly available.

## Acknowledgements

We are grateful to everyone involved in running this project including representatives from each company, the catering providers, and the members of staff at every cafeteria.

